# Airflow and air velocity measurements while playing wind instruments, with respect to risk assessment of a SARS-CoV-2 infection

**DOI:** 10.1101/2020.12.17.20248234

**Authors:** Claudia Spahn, Anna Hipp, Bernd Schubert, Marcus Rudolf Axt, Markus Stratmann, Christian Schmölder, Bernhard Richter

## Abstract

Due to airborne transmission of infection with the coronavirus, the question arose as to how high the risk of spreading infectious particles can be while playing a wind instrument.

To contribute to this question and to help clarify the possible risks, we analyzed 14 wind instruments, first qualitative by making airflows visible while playing and second quantitative by measuring air velocities at three distances (1m, 1.5m and 2m) in direction of the instrument’s bell.

Measurements took place with wind instrumentalists of the Bamberg Symphony in their concert hall.

Our findings highlight that while playing all wind instruments no airflow escaping from the instruments – from the bell with brass instruments, from the mouthpiece, keyholes and bell with woodwinds – was measured beyond a distance of 1.5m from the instruments’ bell, regardless of volume, pitch or what was played. With that, air velocity while playing corresponded to the usual value of hall-like rooms, of 0.1 m/s. For air-jet woodwinds, alto flute and piccolo, significant air movements were seen close to their mouthpieces, which escaped directly into the room without passing through the instrument and therefore generating directed air movements.

## Introduction

The coronavirus pandemic has had and continues to have a grave impact on music making, especially concerning the playing of wind instruments and singing. Airborne transmission plays an important role for the spread of the SARS-CoV-2 virus [1]. Thus, forms of musical sound generation that involve breathing are suspected of being risky. In this regard, it seems of great importance to learn more about airflow and air velocity produced by playing wind instruments and singing that could contain infectious droplets or aerosols and spread them in indoor situations.

Up-to-date research looked into air dispersion, while playing wind instruments or singing, using different forms of measuring.

In a recent publication, conducted at the Bauhaus-Universität Weimar [2] the spread of breathing air while playing wind instruments and singing was observed, using the Schlieren imaging with a Schlieren mirror and the Background Oriented Schlieren method (BOS), as a way to make respiratory air visible. Two professional singers (baritone and soprano) and eleven wind instruments (woodwinds: oboe, bassoon, Bb clarinet, bass clarinet, flute, piccolo, alto flute, and brass: Bb trumpet, tenor trombone, French horn, F tuba) of the philharmonic orchestra Thüringen Philharmonic Gotha – Eisenach, were positioned in front of the Schlieren mirror while playing or singing. The findings show that the spreading range as well as the angle at which the air escapes mouth or outlet, varies strongly from instrument and player, depending on the structure of the instrument, the structure of the mouthpiece, the way an instrument is blown and individual blowing or breathing capacities. In general their measurements with wind instruments reached a maximal distance of 1.12m, measured by side-air movements of the piccolo.

The authors also discovered that special barrier caps – used with brass instruments – do have a significant impact on the spread of air, which can strongly reduce the dispersion, while hardly interfering with the sound of the instrument. Furthermore, they found out that the escaping air ascends due to natural convection or mixes with the surrounding room air [3]. A study by the Ludwig-Maximilians-Universität München and the University of Erlangen [4], pre-published in July 2020, investigated different forms of speaking and singing with 10 professional singers of the Bavarian Radio Choir. They made respiratory flows visible by exhaling smoke of e-cigarettes (not containing nicotine). Tests were conducted in a shaded room, documenting the exhaled aerosol clouds with the help of a high speed camera and laser light. Comparing different settings (singing text, speaking text and singing without text, once with soft and once with loud phonation) they learned that singing text and speaking text reaches comparable mean distances of dispersion at up to 0.85m, while singing without text reached the lowest values of airflow at 0.63m. Even though the mean measurements of air dispersion stay within a reach of 1m, some singers reached airflow distances of up to 1.4m. These measurements were then compared to coughing, which turns out to reach farther distances, with a mean of 1.3m and a maximum of 1.9m. On the base of their study, they announce a suggestion for distance regulations of at least 2m to the front [5].

First insights on their second study on wind instruments, using the same test setting are published online [6]. They point out, that respiratory clouds to the front reach farther than to the side, with instrument-specific outcomes. For an alto flute they recommend 3m spaces to the front and 2m to the side, for all other wind instruments they recommend 2m to the front and 1.5m to the side [7].

Another study conducted by the University of the German Armed Forces in Munich [8], which was pre-published in May 2020, analyzed larger droplets, when singing and speaking, as well as flow-related small droplets when singing and playing wind instruments. The study was conducted with a professional singer, two amateur choir singers, five professional musicians (clarinet, flute, oboe, bassoon, and trumpet) and an amateur brass player (trumpet, trombone, and euphonium). The motion of droplets and air leaving either mouth or outlet was observed during exhalation, which was then illuminated with laser and recorded with a digital camera, producing a series of images that were subsequently quantitatively analyzed. The analysis points out that while singing no more airflow was detectable at a distance of 0.5m, regardless of volume, pitch or if the singer was a professional or an amateur. As for wind instruments the analysis of brass instruments showed strong air movements in front of the instrument, which did not reach farther than 0.5m. Woodwinds produced comparably more airflow, reaching a distance of around 1m. Concluding, they recommend a minimal radial distance of 1.5m between singers and wind instrumentalists [9].

They furthermore point out the risks of infection in choir singing by the fact that there are social contacts between choir members that are usually part of choir repetitions, e.g. during breaks [10].

In addition, there is another study by Parker and Crookston [11] (pre-published in July 2020) which measured aerosols while playing brass instruments and singing. The authors analyzed 7 brass instruments (cornet, horn, baritone, euphonium, trombone, Eb tuba and Bb tuba), investigating the effect that playing for a more extended period of time has on the release of particles with comparison to singing, breathing and using a special barrier cap. To investigate the particles released, they were size sorted and counted with a six-channel laser particle counter. It is figured out that breathing produces more respiratory droplets than playing, and the authors state that the use of a barrier reduces the release of aerosols by 95%. They also found that within variation of time the production of aerosols increases again and fresh air conditioning is required [12].

At the University of Minnesota a study on aerosol generation from different wind instruments analyzed 15 musicians from the Minnesota Orchestra [13] (playing trumpet, bass trombone, French horn, tuba, piccolo, bassoon, oboe, clarinet and bass clarinet) while playing their instruments, breathing and speaking. By use of an aerodynamic particle sizer, the aerosol concentration and size was measured. In comparison to the aerosol generation of speaking and breathing, the instruments are thereafter categorized into low, intermediate and high-risk levels. They point out that air-jet instruments (piccolo and flute) produce aerosols at the outlet and the other half near the embouchure. Bassoon also produces aerosols at the keyholes, as well as the bell. For woodwinds, in general, the mouthpiece and tube structure play a significant role in the generation of aerosols. For brass instruments they find, that the total length of the tube correlates with the concentration of aerosols (trumpet > bass trombone > French horn > tuba) [14].

Furthermore, Mürbe et al. [15] pre-published a study on the increase of aerosols during professional singing. Testing 8 professional singers (two female soprano, two female alto, two male baritone and two male tenor) of the RIAS chamber choir Berlin, they measured particle emission rates with the help of a laser particle counter, during breathing, reading, singing and holding a long tone. They confirm their assumption of singing producing higher emission rates than speaking, with mean measurements of 4.71 – 84.76 P/s during speaking and 753.4 – 6093.14 P/s during singing. Women also produced more particle emission rates than men, leading to the assumption that high voices produce a higher sound pressure level than lower voices. On the base of their findings, they assume that singing produces more emission rates of aerosols than speaking or breathing and that an increase of aerosol rates can be observed with an increasing sound pressure level during singing, especially while holding long tones [16].

In consideration of their study, Mürbe et al. [17], Kriegel and Hartmann [18], and Hartmann, et al. [19] publish various risk assessments on the risk of infection with virus-loaded aerosols, while singing indoors during the SARS-CoV-2-pandemic, assuming that different styles of singing – e.g. singing vs. speaking – as well as different intensity of voice can lead to various sizes and density of droplets and aerosols [20] and that room situations of choir rehearsals have to be taken into account [21, 22].

They also found out that child voices emit fewer aerosols during singing that adults [23]. The study tested 8 children (four girls and four boys) of semiprofessional children’s choirs (Staats-und Domsingknaben Berlin and a girls choir of the Berliner Singakademie), who were all 13 years old (except one girl was 15 years old). The study was conducted the same as their previous study on professional adult singers, as mentioned above. Their mean measurements show emission rates of 16 – 267 P/s for speaking, 141 – 1240 P/s for singing, and 683 – 4332 P/s for shouting [24].

These up-to-date studies looked into the spreading of air or aerosols, while playing wind instruments or singing, focusing on different instruments, singing styles or using different forms of measuring and making airflows visible. They give similar results focusing the visualization of airflows while playing wind instruments and singing. Accordingly different methods were used like the Schlieren method to make respiratory airflows visible [25], the visualization of airborne transmission of professional singers and wind instruments by use of e-cigarette smoke [26, 27], the observation of large and small droplets from wind instruments and singers, on the basis of illuminating airflows with laser and analyzing picture series [28], the release of respiratory aerosols, while speaking and playing brass instruments, as well as for using barrier caps [29], the generation of aerosols of different wind instruments, measured with an aerodynamic particle sizer [30] and the measurement of particle emission rates of professional singers [31] and children [32], leading to several risk assessments of singing indoors during the SARS-CoV-2-pandemic [33, 34, 35].

The measurements of our study took place at the beginning of May 2020, when wind instrumentalists had to maintain distances of 12m following official instructions and recommendations by statutory accident insurance. With this background it was tremendous to bring evidence into the field of music making. In this respect, our study aimed to provide funded data on velocity, direction and distance of respiratory air while playing wind instruments.

## Materials and Methods

### Sample

Players of brass instruments (trumpet, trombone, horn and tuba), as well as woodwind players (alto flute, piccolo, oboe, English horn, clarinet, bass clarinet, bassoon and contrabassoon) of the Bamberg Symphony voluntarily took part in the study. Two further professional musicians – tenor saxophone and recorder players who are not part of the classical orchestra – have also been included, leading to a total sample of 14 wind instrument players.

All persons were asked by the administration of the Bamberg Symphony to take part in the measurements. At this time of lockdown musicians were highly motivated to contribute to research in order to make playing possible again.

Consent for the study was given by the Ethics Committee of the Universitätsklinikum Freiburg.

In order to exclude infectious persons, all players were questioned before the measurements if they had typical symptoms of the Covid-19 disease. Additionally, everyone’s temperature was taken with an electric fever thermometer, directly before entering the concert hall. No one indicated suspicious symptoms and the measurements of the temperature showed values under the cut-off of 37.5°C in all persons.

### Design and procedure

At the beginning of May 2020 measurements were conducted, which were initiated by the Bamberg Symphony Orchestra during the first lockdown of the Covid-19 pandemic in Germany. All solo players for wind instruments of the orchestra (brass: trumpet, trombone, horn and tuba; woodwinds: bassoon, contrabassoon, alto flute, piccolo, oboe, clarinet, bass clarinet and English horn), and an additional professional tenor saxophone and a recorder player, were examined, while playing their instruments.

The measurements took place in a typical surrounding for classical musicians, using the stage of the Bamberg Symphony (see S1 Appendix). The aim of the measurements was to first make respiratory air visible and then to measure air velocities at and coming from the instruments’ bell, as well as other outlets (e.g. keyholes or side airs). We wanted to find out where air velocities can be measured, at what value and to what extent. Therefore, special air velocity sensors were put at the bell of the instrument, at the distances of 1m, 1.5m and 2m (see S2 Appendix, S3 Appendix). For the qualitative analysis, the measurements were also filmed on a digital camera, producing videos. For all wind instruments except recorder, qualitative and quantitative measurements could be analyzed. Due to technical disruptive factors for the recorder only qualitative observations are available.

All wind instrumentalists played scales, excerpts of music pieces and long tones with different pitches and dynamics (pp – ff), as well as different articulations (e.g. staccato). Particular to the instrument, the measurement focused on the escape of air through tone holes and outlets specifically. Since warming up is different for every instrument and concerning brass winds usually includes a lot of blowing through the mouthpiece (without using the whole instrument), these situations were analyzed as well.

#### Visualization of airflow

The flow visualization, by use of artificial mist, is one option for on-site-inspection of airflows. Using the technique of flow visualization with the FlowMaker™, swirls have been qualitatively made visible, at the outlet of wind instruments [36, 37].

Coming into operation was a harmless artificial fog of SAFEX®-Chemie GmbH [38], which consists of water droplets and is usually used as stage fog [39]. The fog droplets have a size smaller 5µm (see S4 Appendix) and can therefore be compared to the dangerous core droplets of the coronavirus [40].

The artificial fog was transported through a system called “Hydra”, using a flexible tube, to the release spot of the instruments, embouchure area, bell, and key openings. Through the application tube, installed on a stand, the fog escaped into the free space of the room and created a cloud of fog. It was oriented towards the musicians, who placed the outlet of their instrument directly into the cloud (see S5 Appendix, showing the horn player).

The movement of the fog was filmed with a video camera and qualitatively analyzed afterwards.

#### Measurement of air velocity (anemometry)

For the measurement of air velocity, omnidirectional (independent of direction) hot film probes type DISA 54N50 Low Velocity Air Flow Analyzer, manufactured by DANTEC were used. The probes have a measuring area of 0 - 1 m/s with an accuracy of 0.2 – 0.4 % full scale. The corresponding electronics (LVFA) give a linear voltage signal of 0 – 2 Volts, according to its velocity, which is recorded with a 20 bit-AD converter in the computer. Before the measurements, the measurement chain (sensor – electronic measurement equipment – signaling cable – converter – computer) was verified in the company-owned wind tunnel against a laser-Doppler anemometer.

With a distance of 1m, 1.5m and 2m from the exhaust opening, ball tubes were put on stands and placed in a line. All sensors were at a height of 1m above ground and were adjusted according to the different instruments and their outlet holes (see S3 Appendix).

The linear output signal of the controller was mapped with a 20 bit AD converter and recorded on the cable-connected computer every second.

The relation of the measuring signal and the actual velocity was verified according to the measurements of the in-house calibration wind tunnel, in the area of 0.15 – 0.7 m/s.

Parallel to the measurements of the velocity, video recordings with a manual camera were also conducted.

The quantitative measurements were used to support the qualitative observations by focusing the distances of 1m, 1.5m and 2m, measuring air velocities in direction of the instruments’ outlets. The video sequences of the qualitative observations were timed with the measurements of the velocity measurement probes, to get the relation of distance, direction and velocity of the emitted air. Hereinafter, the findings are presented by selected measuring charts (e.g. see S6 Appendix). These charts were used as basis for the analysis and were compared to data numbers and video sequences, in order to understand what air velocities were measured where and while doing what kind of playing or warming up.

#### Measurement report

Among circles of experts on air technology, room air velocities are part of indoor climate discussions. The indoor climate of habitable rooms is called comfort climate: “A climate of comfort persists, when people feel thermally at ease in habitable rooms” [41]. Whether a person is feeling comfortable within a room is – amongst other components, like temperature, etc. – dependent of air velocities, since a comfortable climate is free from draught. Draught is the undesired cooling of the body through air movement. It can be felt from a merit of 0.15 m/s, and depends on the size of a room and its ventilation system [42].

The perception of comfortableness of concert halls is therefore closely related to ventilation systems of the hall. So-called “well-like ventilations”, coming from the ground, are usually used nowadays [43]. In this relation, the velocity, in which the air exits the ventilation system has to be taken into account, since it is regulating how comfortable the audience feels. For concert halls exit velocities of 0.2 m/s are recommended (in relation to a room temperature of 20°), to not surpass an air velocity of 0.15 m/s at the height of 1m (were the audience is sitting) and to stay in the comfort zone of 0.1 – 0.2 m/s [44]. Hence, for concert halls, which have a room temperature of 20°, room air velocities of 0.1 – 0.16 m/s are usually estimated [45].

Looking at the circumstances of the concert hall of the Bamberg Symphony, the ventilation system was analyzed in 2017 to understand draught appearances on stage, giving our study funded data on the room air conditions, including the room temperature of 22.3°. Hence, the draught risks – where people start to feel uncomfortable – was stated at 0.15 m/s the highest and therefore considered an area of comfortableness at 0.1 – 0.15 m/s [46].

These numbers show that measurements under a merit of 0.1 m/s cannot be recognized by people and are comparable to “background noises”. Simultaneously, they point out that a merit of 0.15 m/s is already felt as draught, leading to a range of room comfortableness between 0.1 – 0.15 m/s. These numbers help to understand the measurements of the air velocities of wind instruments, giving them a relation and comparability. Merits under 0.1 m/s are therefore not considered, while merits of 0.15 or even 0.2 m/s can be understood as remarkable.

As a basis for the analysis graphs for every instrument were produced, indicating the findings of an instrument. These charts compare the airflow visualizations of the video sequences to the measured numbers of airflows within the three distances (1m, 1.5m, 2m) in direction of the instruments ′ outlet (see S6 Appendix, Appendix S7).

Making use of the descriptive analysis, the measurements of every instrument were also put into a table, comparing air velocities of different instruments, at the three different distances. This overview showed the maximum values of every instrument, at every distance – which were compared and put into a relation to the numbers of room comfortableness. Air velocities produced by a wind instrument that are lying under a value of 0.1 m/s do not have an impact on the compartment air and “disappear” amongst “background noises”, meaning velocities. Whereas air velocities with a value over 0.3 m/s are comparable to strong draughts or coughing and therefore have a strong impact on dispersion of air, and in the following, on the dispersion of core droplets, such as SARS-CoV-2 virus droplets.

## Results

### Qualitative flow visualizations by use of artificial fog

The qualitative observations served the analysis of airflows while playing (see S8 Appendix) and gave a first insight into the movement of airflows from the different outlets of the instruments. Aside from that, these observations were significant to the position of the sensor for the air velocity measurements, showing precisely at what spot of the instrument airflows escaped the instrument. The sensor of the tuba, for example, was positioned above the instrument, since its outlet directs upgrade, whereas the outlet of the oboe points to the floor and the output of the horn points backwards. The main outlet for the observed brass instruments (trumpet, trombone, horn and tuba) was the bell, whereas for woodwinds keyholes as well as airflows close to the mouthpiece have to be considered.

#### Brass

Next to airflows, which escaped at the bell of the brass instruments, further air movements were seen while deflating the instrument, which concerned mainly the trumpet and trombone. This procedure is part of warming-up and generates visible airflows, which can only be seen within a distance of 1m. Comparing these air movements to directed blowing (in direction of the sensors), blowing without the instrument showed stronger and faster air movements in the artificial fog. While the trumpet and trombone players were playing an excerpt of a music piece, only very small air movements were made visible, which mixed with the surrounding room air velocity quickly. As for the tuba and horn no air movements were made visible, while playing.

#### Air-jet woodwinds

For the piccolo and alto flute side airflows could be made visible, escaping the mouth close to the mouthpiece, directly reaching to the ground and staying visibly close to the player’s body. Aside from these significant observations no further airflows could be seen at the bell of these two air-jet instruments.

For the recorder (early baroque in g, and soprano) small air movements were visible at the labium of the instrument, at a distance of max. 20 cm (see S9 Appendix). Further airflows were not visible at the bell of the recorder, leading to the presumption that the labium can be understood as the instruments’ main outlet (see S10 Appendix).

#### Reed woodwinds

Further reed woodwind instruments, which were measured (single reeds: clarinet, bass clarinet, saxophone and double reeds: bassoon, contra bassoon, oboe and English horn) mainly showed airflows escaping the instruments’ bell, with little observations at the keyholes. Even though these instruments showed airflows escaping from the keyholes as well, these air movements showed only little dispersions in the artificial fog.

While playing the clarinet, for example, small airflows were visible at the bell of the instrument, which were thereafter compared to blowing without the instrument. This comparison showed how small the air movements with the instrument were, in comparison to the fast and strong airflows, produced by blowing from the mouth of the clarinetist without the instrument.

### Measurements of air velocity (anemometry)

The air velocity measurements with all wind instruments mostly did not surpass a value of 0.1 m/s (see Table 1), while playing an excerpt of a music piece, scales, or different pitches and volumes. Even though air movements were qualitatively seen at the bell of some wind instruments, these qualitative observations did not reach measurements of more than 0.1 m/s, which is the value of usual room air velocities in hall-like rooms. Hence, some little air movements were measurable at the 1m sensor (concerning tuba: 0.13 m/s, oboe: 0.15 m/s and contrabassoon: 0.11 m/s – at the 1m sensor), not surpassing measurements of 0.1 m/s at the 1.5 or 2m sensor.

**Table 1.** Maximum measurements of all test instruments, while playing an excerpt from a music piece:

| Instruments | 1m | 1.5m | 2m |
| --- | --- | --- | --- |
| <u>brass</u> |  |  |  |
| trumpet | <0.1m/s | <0.1m/s | <0.1m/s |
| trombone | <0.1m/s | <0.1m/s | <0.1m/s |
| horn | <0.1m/s | <0.1m/s | <0.1m/s |
| tuba | 0.13m/s | <0.1m/s | <0.1m/s |
| <u>woodwinds</u> |  |  |  |
| 1) air-jet |  |  |  |
| alto flute | <0.1m/s | <0.1m/s | <0.1m/s |
| piccolo | <0.1m/s | <0.1m/s | <0.1m/s |
| 2) single reed |  |  |  |
| clarinet | <0.1m/s | <0.1m/s | <0.1m/s |
| bass clarinet | <0.1m/s | <0.1m/s | <0.1m/s |
| saxophone | <0.1m/s | <0.1m/s | <0.1m/s |
| 3) double reed |  |  |  |
| oboe | 0.15m/s | 0.12m/s | <0.1m/s |
| bassoon | <0.1m/s | <0.1m/s | <0.1m/s |
| contrabassoon | 0.1m/s | <0.1m/s | <0.1m/s |
| English horn | <0.1m/s | <0.1m/s | <0.1m/s |

### Brass instruments

The playing of trumpet, trombone, horn and tuba showed only little or no airflows in the artificial fog. However, there was a difference between playing and warming up. Since brass players often use only mouthpieces while warming up, direct airflows are produced, which can reach measurements higher than 0.1 m/s. In comparison, loudness or pitch did not have an impact on the air velocities (see S6 Appendix, S7 Appendix).

#### Trumpet

While playing different scales, high and medium pitches, all measurements of the trumpet stayed under 0.1 m/s at any distance, which corresponds with the usual room air velocity.

However, when the player deflated the instrument air velocities reached 0.14 m/s at a distance of 1m from the bell, while at 1.5m and 2m they stayed under 0.1 m/s. Comparing these numbers to the values of blowing without the instrument, the measurements were much higher, producing air movements of 0.52 m/s to the front, at a distance of 1m. At 1.5m during blowing without the instrument they dropped to 0.15 m/s and at a distance of 2m they stayed under 0.1 m/s.

#### Trombone

Similar to the measurements of the trumpet, air velocities of the trombone did not surpass a value of 0.1 m/s while playing scales and different pitches, and even while deflating, at any distance.

When the person only blowing, without the instrument or the mouthpiece, the highest measurements were made, reaching a value of 0.4 m/s at 1m, 0.18 m/s at 1.5m to the front and 0.1 m/s at 2m.

#### Horn

For the horn all measurements stayed under a value of 0.1 m/s at all three distances, regardless of whether they were playing an excerpt from a music piece, different pitches or volumes.

#### Tuba

The tuba did reach the highest measurements of all brass instruments during playing a music piece or warm up with the instrument. While playing scales and while deflating they did not surpass a value of 0.1 m/s, at every distance. But, when the player did an excerpt of a music piece or warmed up the instrument, the measurements reached 0.13 m/s at a distance of 1m upwards (in the direction of the bell), and stayed again under 0.1 m/s at 1.5m and 2m.

### Wood-wind instruments

For some wood-winds, the measurements indicate that air movements were not only visible at the bell of the instrument, but also at other outlets, such as side-air observations or airflows escaping tone holes – concerning difference between air-jet woodwinds and reed woodwinds. As for single reed and double reed woodwinds, there were no significant differences in the measurements.

#### Air-jet woodwinds

Since there were qualitative observations of side-air movements for both air-jet woodwinds, another sensor was put up at 0.5m to the side of the player, measuring side-air velocities.

#### Alto flute

Regardless of what the flute player was playing all measurements to all sides and distances stayed under 0.1 m/s. The additional measurements to the side reached a value of 0.15 m/s at a distance of 0.5m.

#### Piccolo

The measurements at the bell of the piccolo stayed under a maximal value of 0.1 m/s and therefore mixed with the usual room air velocity of the concert hall. Other than the flute, side-air measurements for the piccolo showed a slightly smaller value of 0.13 m/s.

#### Single reed woodwinds

The qualitative observations showed air movements at the keyholes of several reed woodwinds, which were considered due to the front sensor at 1m.

#### Clarinet

The measurements for the clarinet, at all distances did not surpass a value of 0.1 m/s, regardless of whether they were playing scales, different pitches or an excerpt from a music piece.

#### Bass clarinet

Air velocity measurements for the bass clarinet did not surpass a value of 0.1 m/s, with no difference of what was played (an excerpt from a music piece, long tones or staccato), at a distance of 1m in the direction of the bell, and stayed under a value of 0.1 m/s at 1.5m and 2m.

#### Tenor saxophone

The measurements for tenor saxophone playing did not reach a measurement of air velocity over 0.1 m/s at all distances – regardless of what the player was playing.

#### Double reed woodwinds

##### Oboe

The measurements of the oboe reached a value of 0.15 m/s at a distance of 1m, 0.12 m/s at 1.5m and stayed under the usual room air movement of 0.1 m/s at 2m.

##### Bassoon

All measurements of the bassoon stayed under a value of 0.1 m/s at all distances, independently of what was played.

##### Contrabassoon

All measured data of the contrabassoon stayed at 0.1 m/s, with no difference between long tones or an excerpt from a music piece, at all distances from the bell.

##### English horn

While playing the English horn, regardless of what was played, all measurements in direction of the bell stayed under a value of 0.1 m/s.

### Woodwind and brass instruments, with ambient noises

As a counter phase to the individual measurements, situations with ambient noises were also considered, since they happen frequently during orchestra rehearsals or concerts, e.g. while setting up, rebuilding the stage or simply while everyone walks to their seats. Within these test set-ups the musicians were also playing, while someone walked across the room or other people were talking to each other.

During these measurements we tried to differentiate between air velocities coming from the players or the surroundings. Hence, it was shown that the air velocities could clearly be separated, since they stayed very close to the instrument’s player (see S11 Appendix) or came from the disturbing noises, which rose instantly as soon as someone passed the measurement points or spoke to the player. Simply raising a hand making a very small movement did show air velocity measurements at the closest sensor, making it very sensitive to airflow movements.

The measurements concerning ambient noises are obviously higher than those of measuring individual instrumentalists. They rose up to 0.55 m/s, when people around the players were talking and stayed within a measurement range of 0.18 – 0.55 m/s, when someone walked pass the person playing.

## Discussion

This study observed 14 wind instruments played by soloists of the Bamberg Symphony: trumpet, trombone, horn, tuba, alto flute, piccolo, oboe, clarinet, bass clarinet, bassoon, contra bassoon, English horn, saxophone and recorder (the last two instruments by external professional players). Since the measurements with all players were conducted in the concert hall of the Bamberg Symphony, it has a high external validity for professional classical music settings.

On the basis of our air velocity measurements, we find that distance regulations of 2m to the front and 1.5m to the side are maintainable. A finding that is supported by Becher, et al. [47], Kähler and Hain [48], He, et al. [49], Echternach, et al. [50], and Mürbe, et al [51]. The values of the analyzed wind instruments mostly did not surpass a value of 0.1 m/s at all distances while playing an excerpt from a music piece, scales, or different pitches and volumes, with exceptions for the tuba: reaching a value of 0.13 m/s at 1m, oboe: 0.15 m/s at 1m and contrabassoon: 0.11 m/s at 1m. These exceptions concern measurements at the 1m sensor, all of them not surpassing measurements of 0.15 m/s, which is still considered to be part of the comfortable room air climate [52]. Furthermore, some slight air movements were qualitatively seen at the bell of some wind instruments. These qualitative observations did not reach measurements of more than 0.1 m/s, which is the value of usual room air velocities in hall-like rooms.

Only one study, conducted at the LMU [53] suggests a farther distance regulation of 3m to the front and 2m to the side for alto flute players, since the investigators observed farther respiratory air clouds for this instrument especially. On the basis of our observations as well as the measurements, air-jet woodwinds produce strong side air movements, which did, in our case, stay within a reach of 1m to the player. Therefore, we cannot relate to the suggestion of 3m to the front and 2m to the side, but agree with the fact that side air movements have to be seriously considered.

Due to their production of side air movements, the air-jet woodwinds alto flute and piccolo receive a special position in our findings. They reached high measurements of 0.13 m/s (piccolo) and 0.15 m/s (alto flute) to the side at a distance of 0.5m, which escaped directly into the room. This observation highlights the importance of the structure and the mouthpiece of wind instruments and that it is relevant which wind instrument is taken into account. Becher et al. [54] support this finding, by observing a maximal value of air dispersion at 1.12m from the mouth of the piccolo player into the room. The measurement from the bell of the piccolo though was around 0.2m. The high number of 1.12m came from the side air, which was released due to way the instrument is overblown. This finding also corresponds with Becher et al.’s [55] finding on the importance of individual blowing techniques for air movements. We thus affiliate with Becher et al.’s [56] as well as He et al.’s [57] assumption that the structure on an instrument as well as the way a mouthpiece is blown has a significant influence on the generated air velocity while playing, the distance it reaches, and in the following, its generation of aerosols [58].

Hence, our findings do point out that there is a difference between playing the instrument and warming up the instrument, by simply using a mouthpiece – a typical practice for brass winds. As with the study of Kähler and Hain [59], we observed that pitch or volume do not have a significant impact on the velocity of air movements. Thereinafter, we found out that using only a mouthpiece for warm-up-playing, produces strong and fast airflows. Measurements of up to 0.5 m/s were made for warming up, in relation to 0.13 m/s for playing (at the 1m sensor), confirming the suggested visual observations. These warm-ups usually take place in single rooms, before the concert, but they are often conducted as well during a concert or rehearsal. Regarding the high air velocities produced we strongly suggest, to not blow through a mouthpiece (without the instrument), when other musicians are around.

Comparing brass and woodwinds, it was qualitatively seen as well as quantitatively measured that professional brass players do not produce air movements at the mouthpiece, but at the bell of the instruments. Unlike Kähler and Hain [60], we did not observe a severe difference between the production of respiratory air movements between woodwinds or brass winds. The difference we observed lies more within the structure of the instrument and its mouthpiece, as mentioned above. He et al. [61] also point out the difference between brass and woodwinds, which we can relate to but not support to its full content, since air velocity measurements did not diversify significantly.

As for woodwinds, a difference between so-called air-jet woodwinds and reed woodwinds was not only qualitatively observed, but also quantitatively measured.

The difference between reed and double reed woodwinds was not significant according to our measurements, and is therefore not taken into account. But it has to be mentioned, that reed woodwinds produce little air movements at their tone holes. These air movements were seen in the qualitative observations, but did not reach significant values that would surpass usual room air velocities of 0.1 m/s.

As for the measurements of the oboe, which are comparably high (0.15 m/s at 1m and 0.12 m/s at 1.5m), we assume that surrounding air velocities lead to these values. Otherwise, the values are not plausible, considering the way the instrument and the mouthpiece are played and also regarding our qualitative observation. It can be expected that the measurements should be similar to those of other double reed instruments, such as bassoon or contrabassoon, which reach a merit of 0.1 m/s at 1m (for the contrabassoon) and stay below 0.1 m/s beyond 1.5m (for both double reeds).

Aside from our findings, some limitations of our study have to be taken into account.

First, the study was conducted with highly professional classical wind instrumentalists and the results can therefore not automatically be taken into account for other musical genres and settings or amateur musicians.

Second, the test situation was rather specific. We took into account different test settings: playing setting up the stage (concerning noises and movements of the surrounding) and playing with nothing else happening around. Furthermore, the difference between playing and warming-up was considered. Therefore, the findings are very representative for orchestras and a high level of playing, with restricted transferability for the amateur music sector. Since the players have been a part of the analysis and know their instruments very well, relevant airflow outputs were identified, for each instrument. This seems to be one of the strengths of our study.

Further limitations concern the fact that the measurements were conducted with one person per instrument only, while being sensitive to individual differences of blowing or lung volume, etc. And they were only performed once for every instrument, whereas more repetitions of the same sequence played, would have given more information on reproducibility of the test setting.

Another limitation for the measurement is the fact that the air velocity measurements are very sensitive to surrounding movements, with the waving of a hand already influencing the measurements at the sensors.

## Conclusion

The test results have pointed out that most wind instruments do not have any visual or measureable influence on the movement of compartment air. Merely while playing alto flute light flow movements were visible, close to the musician’s body. Regarding all wind instruments, beyond a distance of 1.5m to the front, no airflows could be measured and therefore no difference compared to usual airflows of hall-like rooms (e.g. movie theatres, theatres, auditoriums, opera, etc.) could be located.

Since no respiratory air movements – of any wind instrument analyzed – were measured at the 2m sensor, we find distance regulations of 2m to the front of wind instrument players maintainable.

In order to maintain a responsible risk management, we find it crucial that besides distance regulations and line-up (e.g. large ensembles), constant fresh air conditioning and the social behavior are being considered. Starting from April 2020, the Freiburg Institute of Musicians’ Medicine constantly updated an official paper on “risk assessment of a coronavirus infection in the field of music” (updated: 2020 May 19, 2020 July 1, 2020 July 17, 2020 December 14), stating the findings of our various studies publicly [62] and establishing a permanent consultation on questions concerning the relationship between the coronavirus and music making.

## Data Availability

All data can be requested by the corresponding author.

## Acknowledgements

We thank all musicians who voluntarily took part in the study.

## Supporting Information

**S1:**
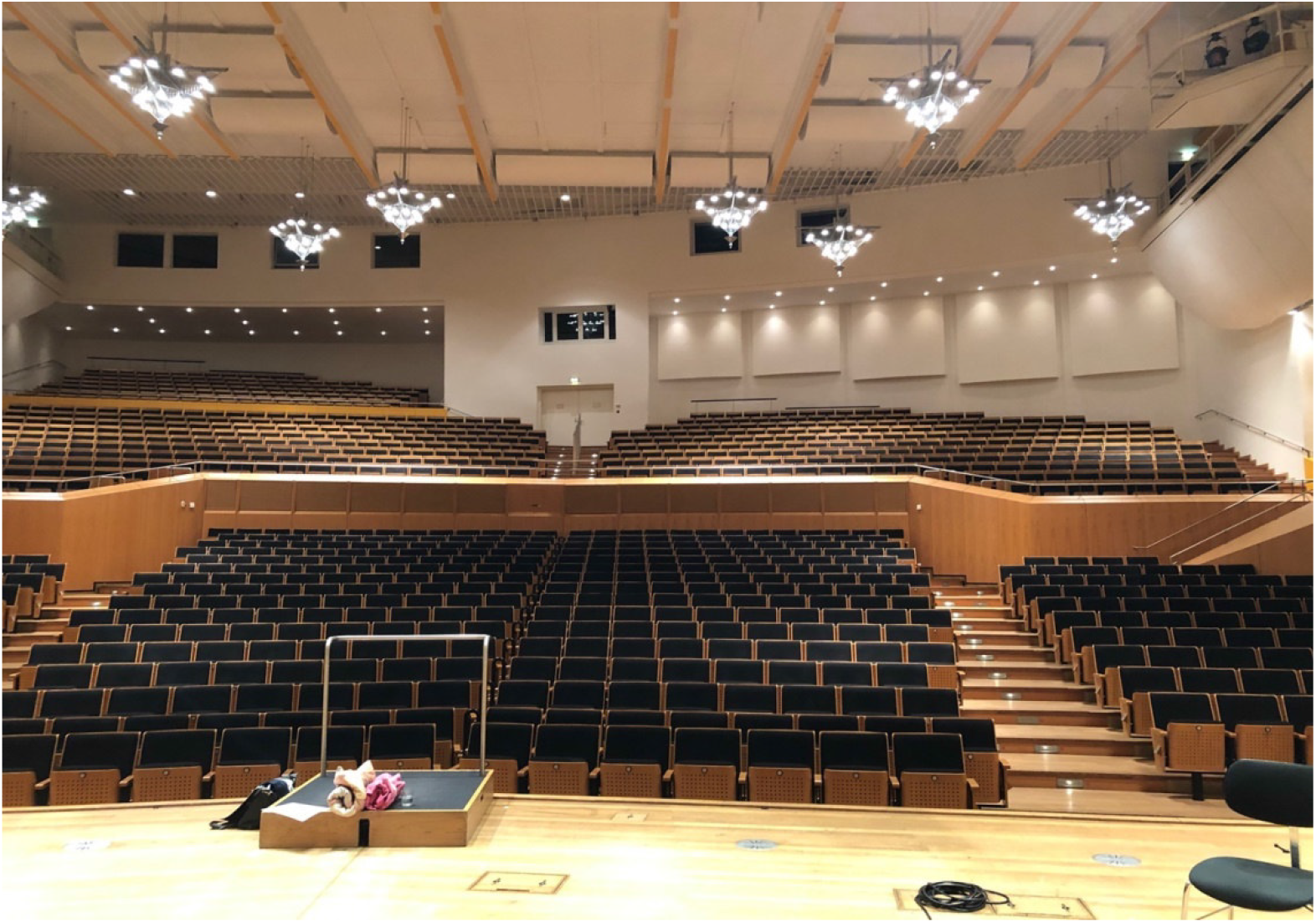
Test location: Concert hall of the Bamberg Symphony

**S2:**
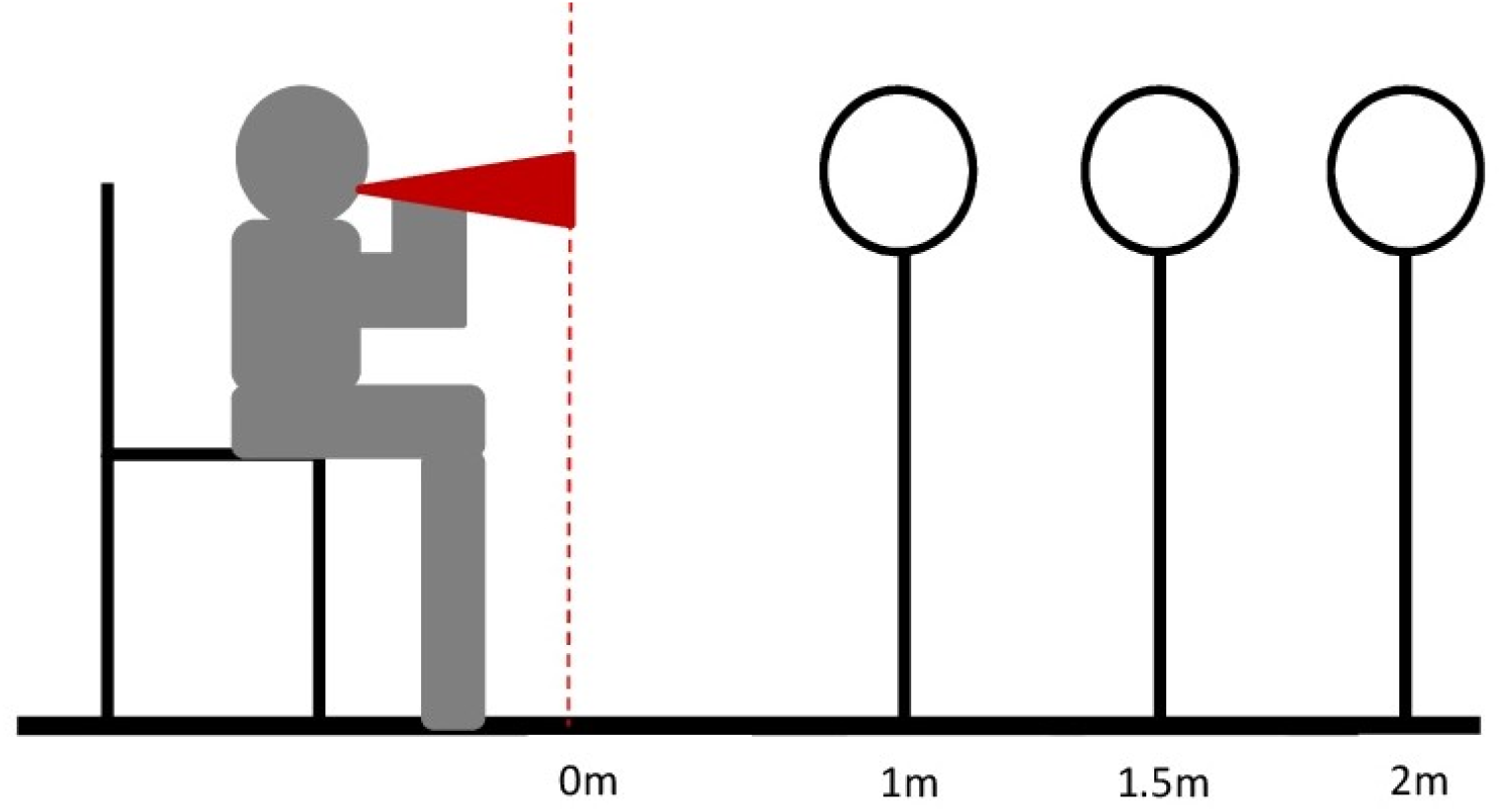
Distance measurements with three sensors in direction of instruments ′ bell

**S3:**
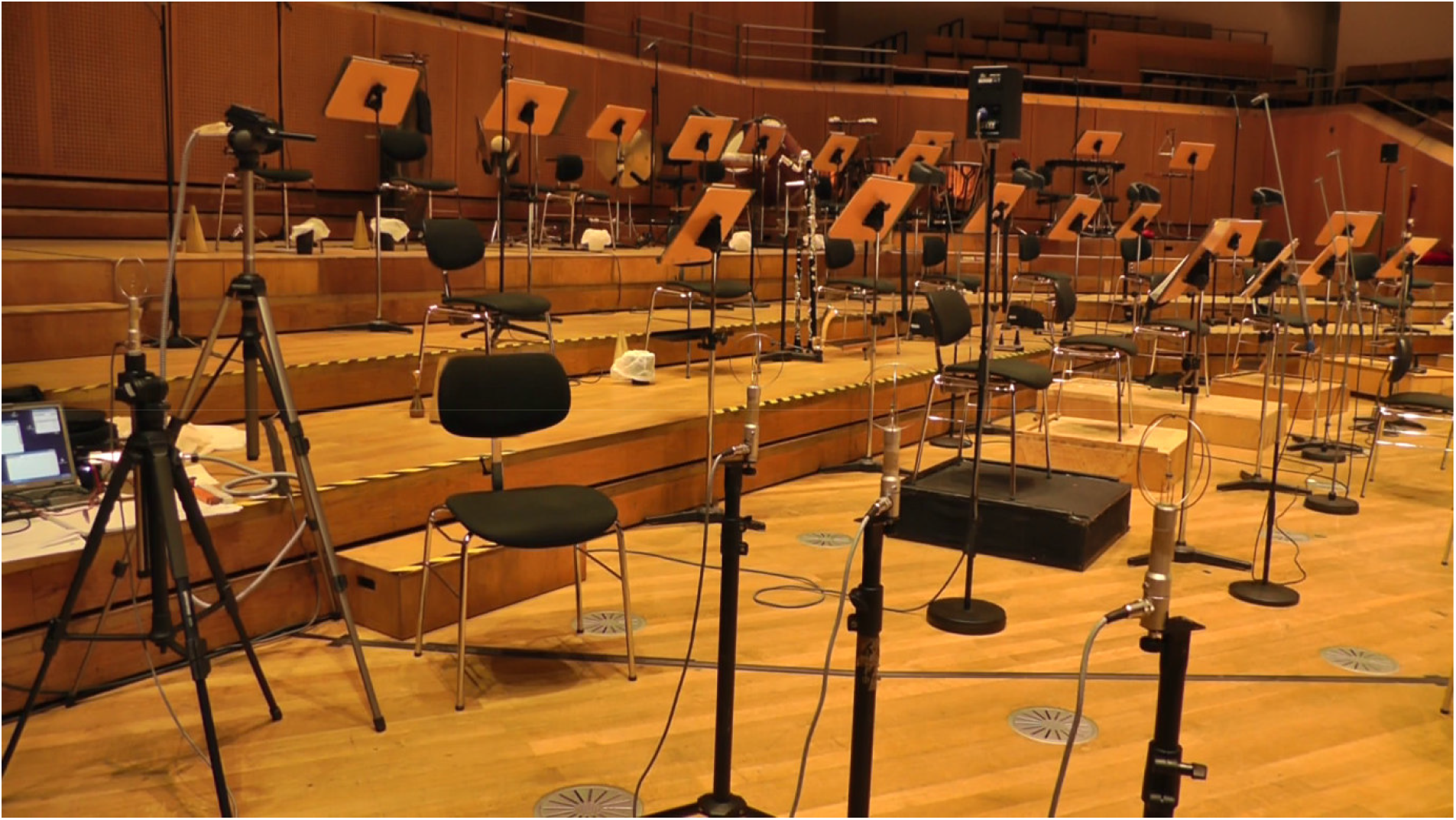
Test set-up on stage of the Bamberg Symphony hall

**S4:**
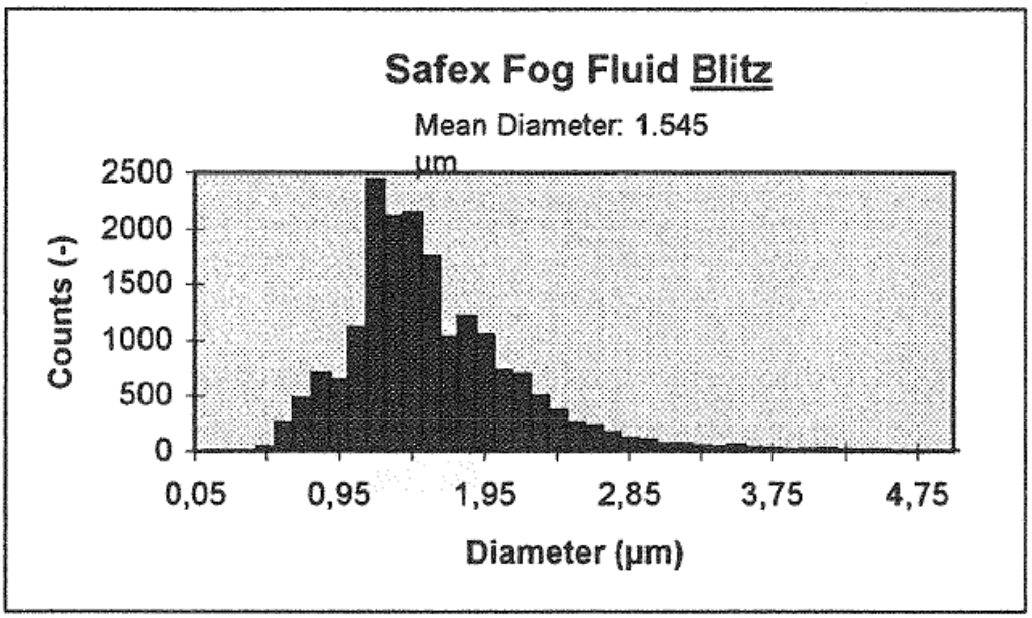
Size distribution of fog droplets (measurements with DANTEC PDA)

**S5:**
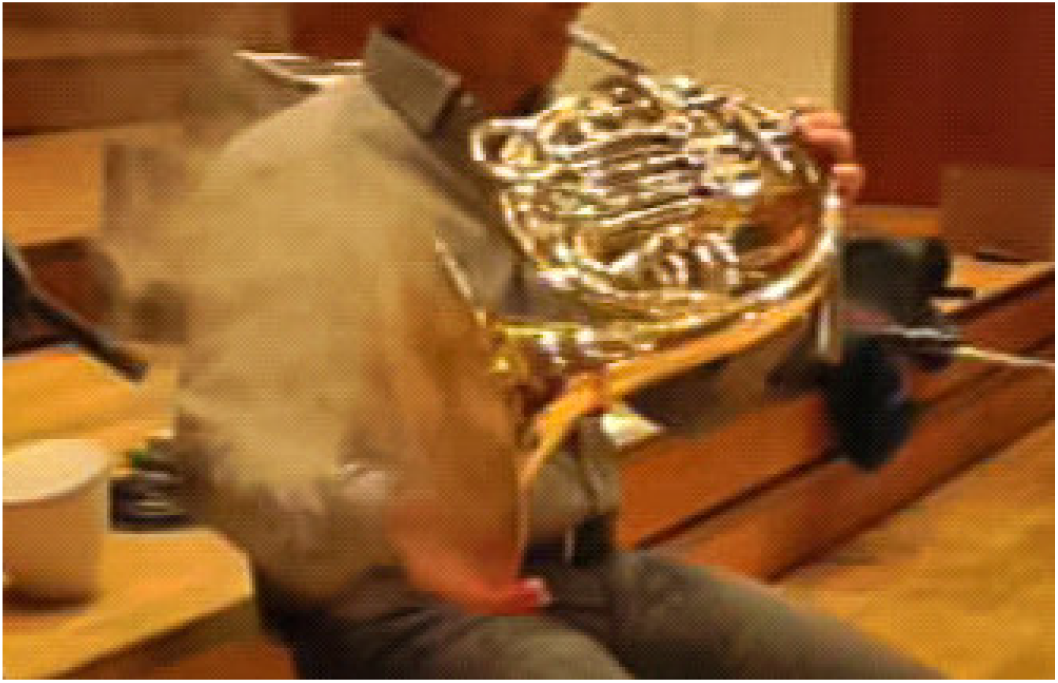
Artificial fog at the bell of the horn

**S6:**
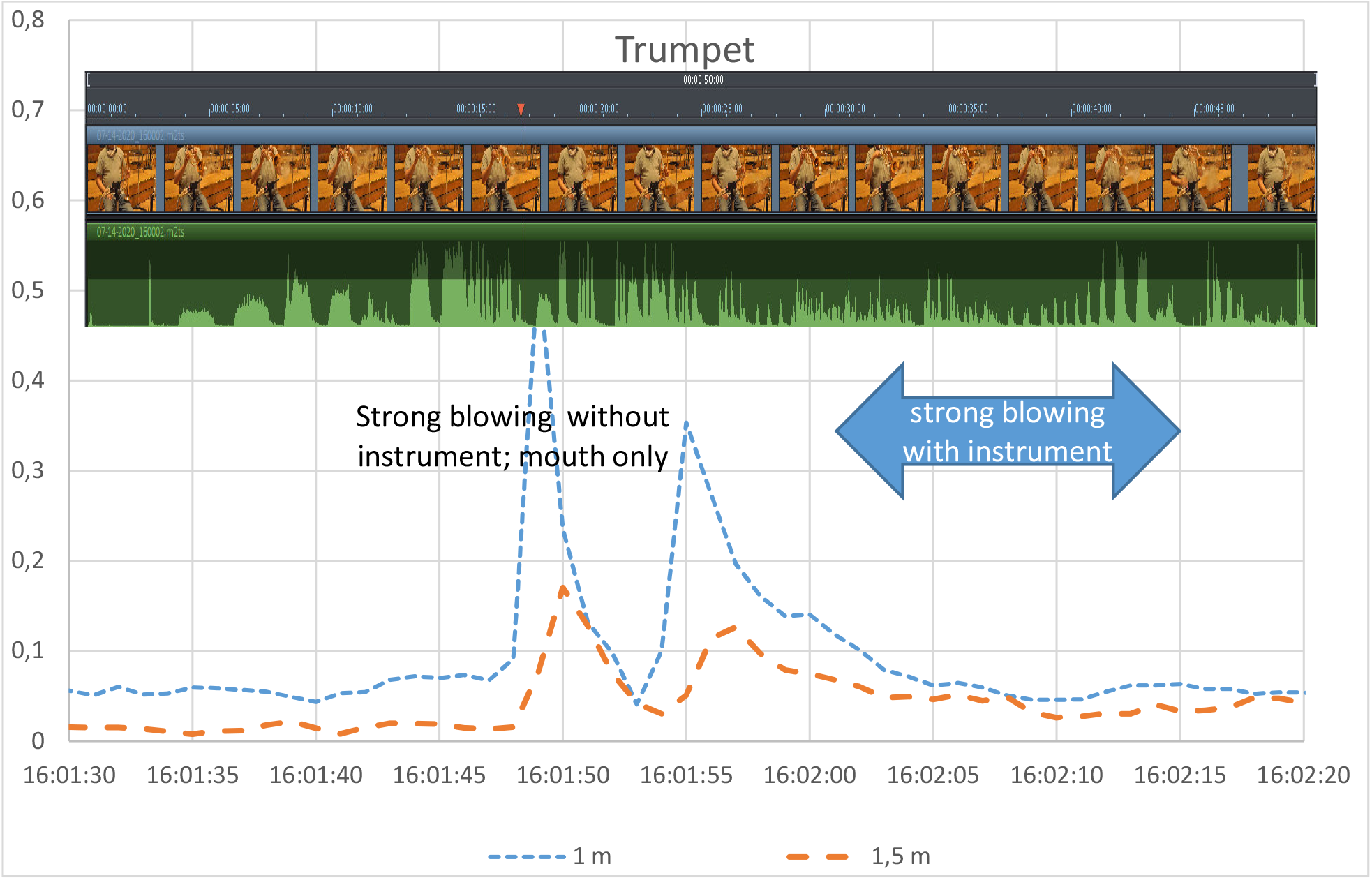
Measurements trumpet: blowing, with and without instrument

**S7:**
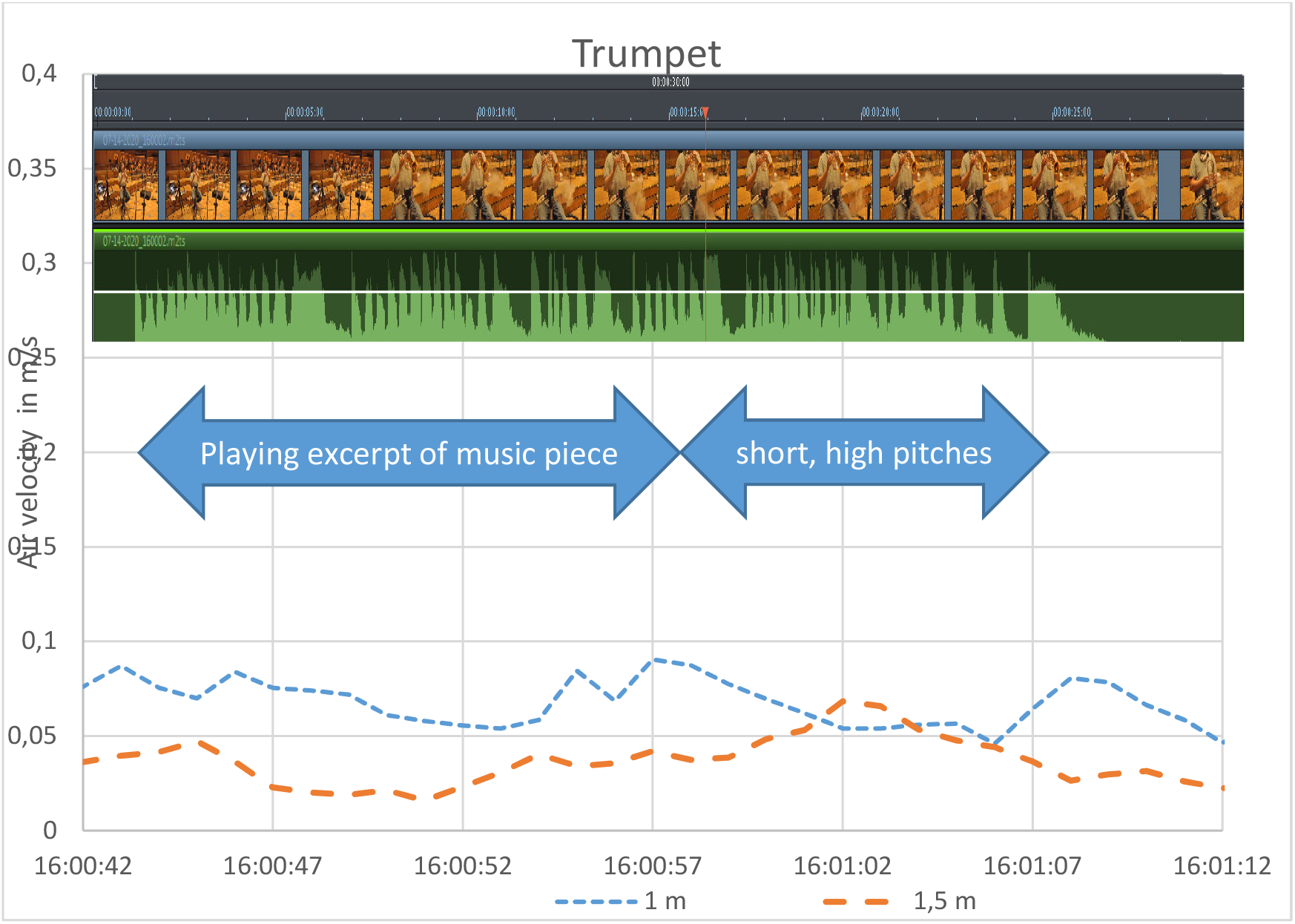
Measurements trumpet: playing and pitches

**S8:**
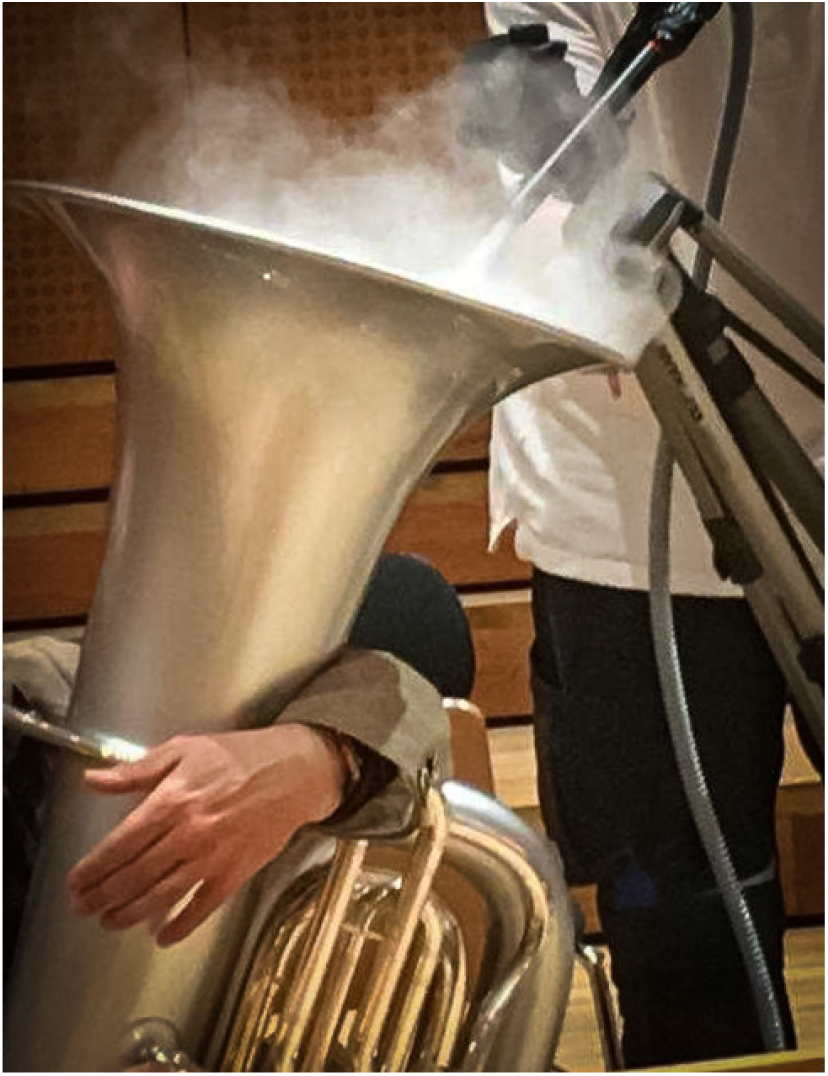
Cloud of artificial fog at the bell of the tuba

**S9:**
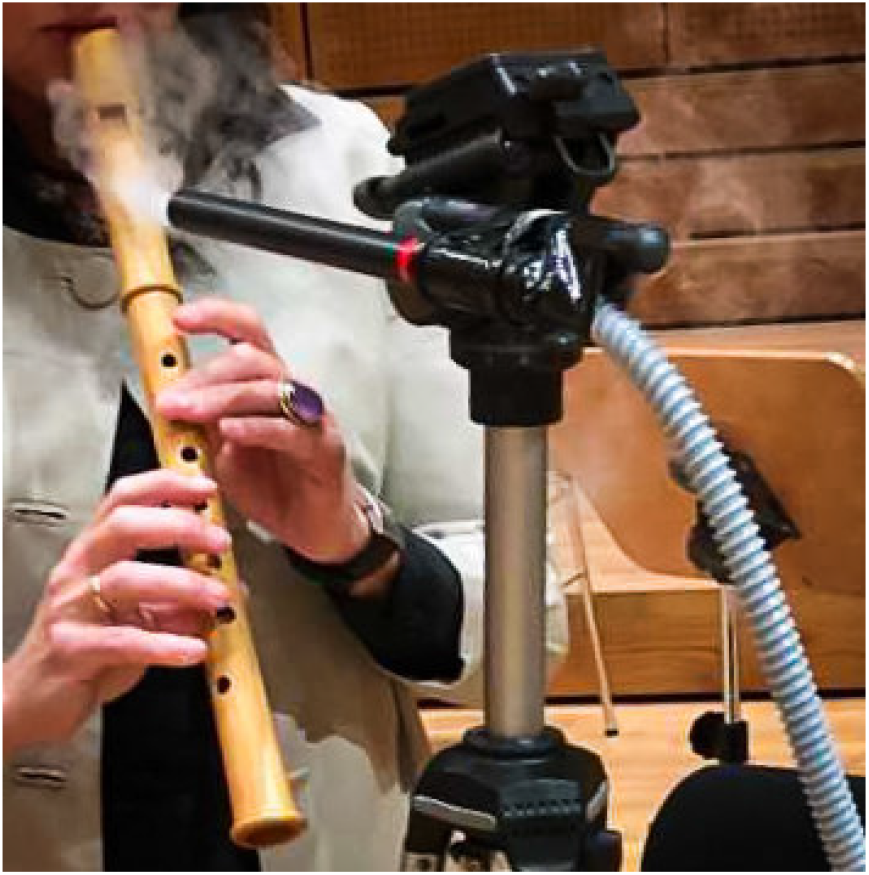
Recorder player while playing, with artificial fog at labium

**S10:**
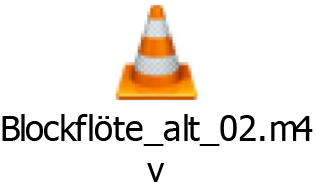
Video for G recorder, while playing, with artificial fog showing airflows

**S11:**
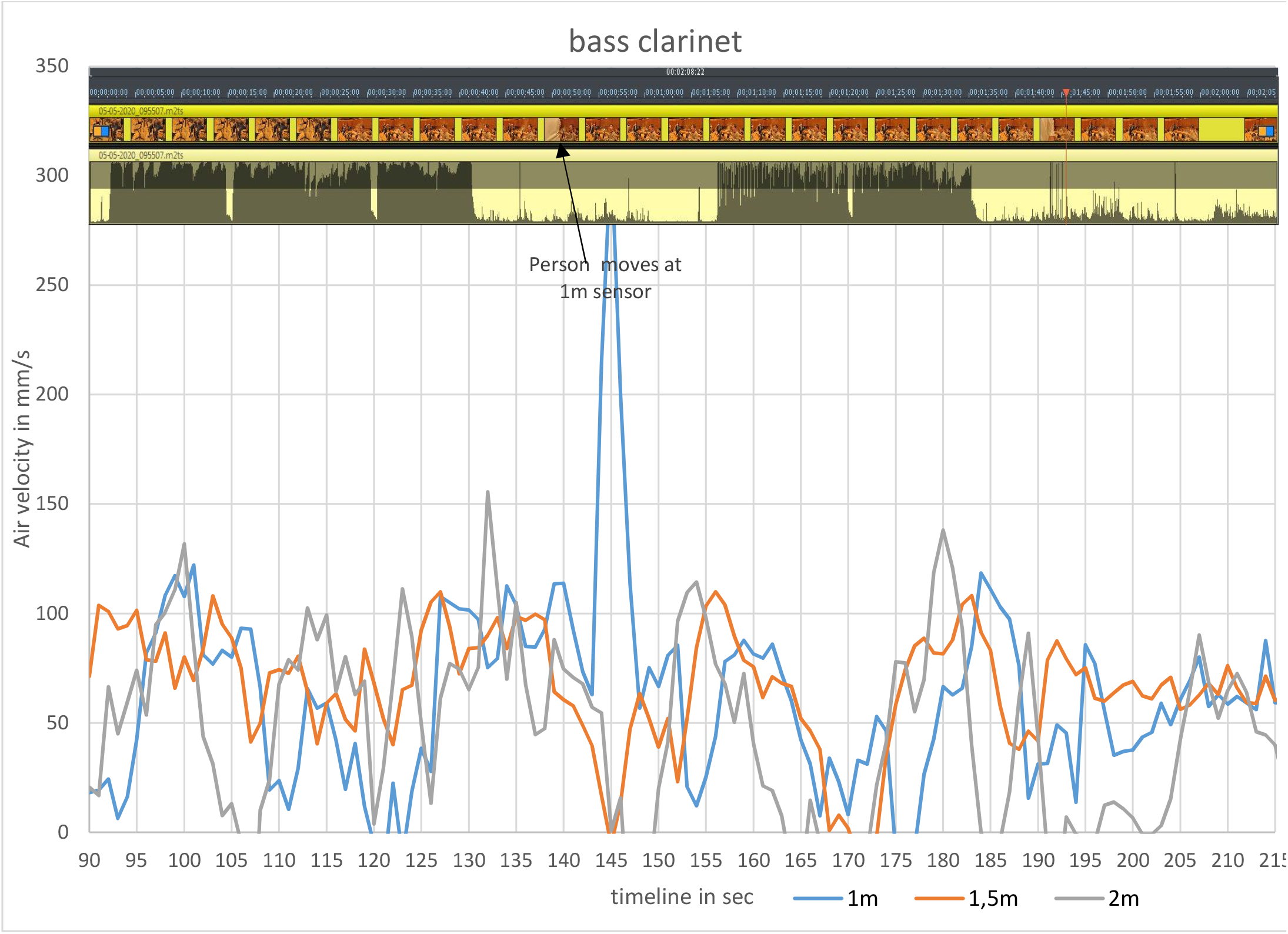
Measurements for bass clarinet, with impact of person passing at sensor

